# Does your measure matter? A comparison of alcohol availability measures in Great Britain

**DOI:** 10.1101/2022.08.18.22278948

**Authors:** Colin Angus, John Holmes, Robin Purshouse, Petra Meier, Alan Brennan

**Affiliations:** School of Health and Related Research, University of Sheffield, UK; Department of Automatic Control and Systems Engineering, University of Sheffield, UK; MRC/CSO Social and Public Health Sciences Unit, University of Glasgow, UK

## Abstract

**Introduction:** The relationship between the spatial availability of alcohol and alcohol-related harm is well established; however the specific components of availability which drive this relationship and how availability may best be measured remains unclear. This study applies a range of alternative measures of availability to selected areas in Great Britain to establish to what extent they are related to each other.

**Methods:** Using a database giving low-level geographic location (full postcode) and type e.g. (bar, supermarket) of every licensed outlet selling alcohol across 4 times points (2003, 07, 10, 14) we calculated low-level measures of alcohol availability for three Local Authorities covering a spectrum of population density and deprivation. Measures were calculated based proximity, outlet density, outlet clustering and so-called ‘gravity’ measures. The relationships between measures are explored descriptively and through correlation and factor analysis.

**Results:** The availability of alcohol is high in the selected Local Authorities, with an average of 45 outlets within 1km of any individual postcode and a mean distance of 400m to an on-trade and 900m to an off-trade outlet. Different measures display different temporal trends with the density and gravity measures having close agreement with each other while proximity measures suggest different trends. Socioeconomic patterning varies both between Local Authorities and measures chosen, with similar agreement between density and gravity measures and different patterns displayed for proximity measures.

**Conclusions:** These findings highlight that different measures applied to different geographies may give fundamentally different characterisations of availability. The choice of availability measure used in any study does, therefore, matter and should not be made without appropriate regard to theoretical considerations

## Introduction

Controlling the physical availability of alcohol is widely accepted as a key policy approach for the reduction of alcohol-related harm [1]. A substantial body of literature has established significant relationships between the increased spatial availability of alcohol and a wide range of outcomes, including alcohol consumption, assault, drink-driving and alcohol-related mortality [2–4]. In spite of the overall consistency of these findings, concerns have been raised about conflicting results when researchers have sought to identify which specific components of availability (e.g. disaggregating by outlet types – bars, liquor stores etc.) are related to which outcomes [5,6].

A recent review identified that the vast majority of availability studies used measures based on either outlet density (118/136 studies reviewed) or outlet proximity (16/136 studies reviewed) [7]. Density measures are calculated by counting the number of outlets within a given geographical region (e.g. administrative divisions, city blocks) and may be weighted for each region by factors such as physical area (e.g. outlets/km^2^), population (e.g. outlets/capita) or roadway miles. Proximity measures are primarily calculated by taking the distance, either Euclidean or using some measure of travel distance, to the nearest outlet. Two alternative measures have been proposed, although neither has been used widely to date: outlet clustering [8] and retail gravity [9]. Cluster measures use spatial geographic methods to identify agglomerations of outlets (e.g. groups of bars) which may have a particularly strong link to intoxication or criminal behaviour. Finally, gravity measures combine the concepts of density and proximity to measure the overall ‘pull’ on a location of all outlets in the surrounding area.

This dominance in the existing literature of simple density or proximity measures to quantify the accessibility of alcohol may, to some extent, be reflective of the fact that researchers are frequently restricted in the level and detail of data which may be available on alcohol-selling outlet locations and types as well as the geographical disaggregation available in their outcomes of interest. The ways in which availability is measured may therefore be due to considerations of convenience or feasibility, rather than having a strong theoretical or methodological basis. This may be considered problematic in view of suggestions that the choice of measure may affect the study findings [10].

This study uses low-level geographic data to calculate four types of measures of alcohol availability for 3 heterogeneous Local Authorities in England across 4 time points. Density, proximity, cluster and gravity measures are calculated and compared to examine the extent to which they correlate with each other, whether their interrelationship is moderated by population density or socioeconomic deprivation and how sensitive each measure is to changes over time. These comparisons will aid researchers in understanding the implications of their choice of availability measure.

## Methods

### Data

Alcohol outlet data for the years 2003, 2007, 2010 and 2014 was obtained from market research specialists CGA Strategy/ Nielsen. Each year of data consisted of a database containing the outlet type and geographic location at full postcode level (a single UK postcode typically contains 15 postal delivery addresses) for all trading outlets at which alcohol was sold. Outlets are categorised as being either on-trade, where alcohol is sold for consumption on the premises (e.g. pubs, bars and restaurants) or off-trade, where alcohol is sold for consumption elsewhere (e.g. supermarkets and grocery stores). This data is compiled by CGA/Nielsen from a wide range of sources including local licensing boards, major alcohol producers and retailers and third party business directories and is estimated to cover 98% of all existing outlets.

Geospatial data containing the detailed grid reference (maximum resolution 1m) of the address-weighted centroid of each full postcode was obtained from the Office for National Statistics (ONS) [11]. This data was mapped to the outlet data, allocating a physical location (i.e. the relevant postcode centroid) for each outlet. A spatial area (in metres squared) was calculated for every postcode using Postcode boundary polygon data from the ONS.

The most widely-used measure of socioeconomic deprivation in the UK is the Index of Multiple Deprivation (IMD) [12]. This is a composite measure, including income, employment, education, health, crime, housing, access to services and environmental factors, which is calculated at Lower Super Output Area (LSOA) level, with each LSOA typically comprising 400-1,200 households. IMD estimates are calculated approximately every 3 years. For every year of outlet data (2003, 07, 10 and 14) we matched every LSOA to the most recently-calculated IMD measures, derived quintiles of the distribution by rank order and allocated these to the postcodes within each LSOA.

In order to explore the moderating impact which the geographical context may have on the relationship between measures of availability, we selected 3 exemplar Local Authority areas in England which epitomise a range of geographies. Islington is a metropolitan borough in central London; Norfolk, a rural country in the east of England; Sheffield, a post-industrial city in the north of the country. Demographic characteristics of the three areas are shown in Table 1 and Figure 1.

**Table 1.** Area and population of included Local Authorities.

| Local authority | Area (km<sup>2</sup>) | 2014 Population | Population per km<sup>2</sup> |
| --- | --- | --- | --- |
| Islington | 15 | 221,000 | 14,735 |
| Norfolk | 5,380 | 878,000 | 163 |
| Sheffield | 368 | 564,000 | 1,532 |

**Figure 1.**
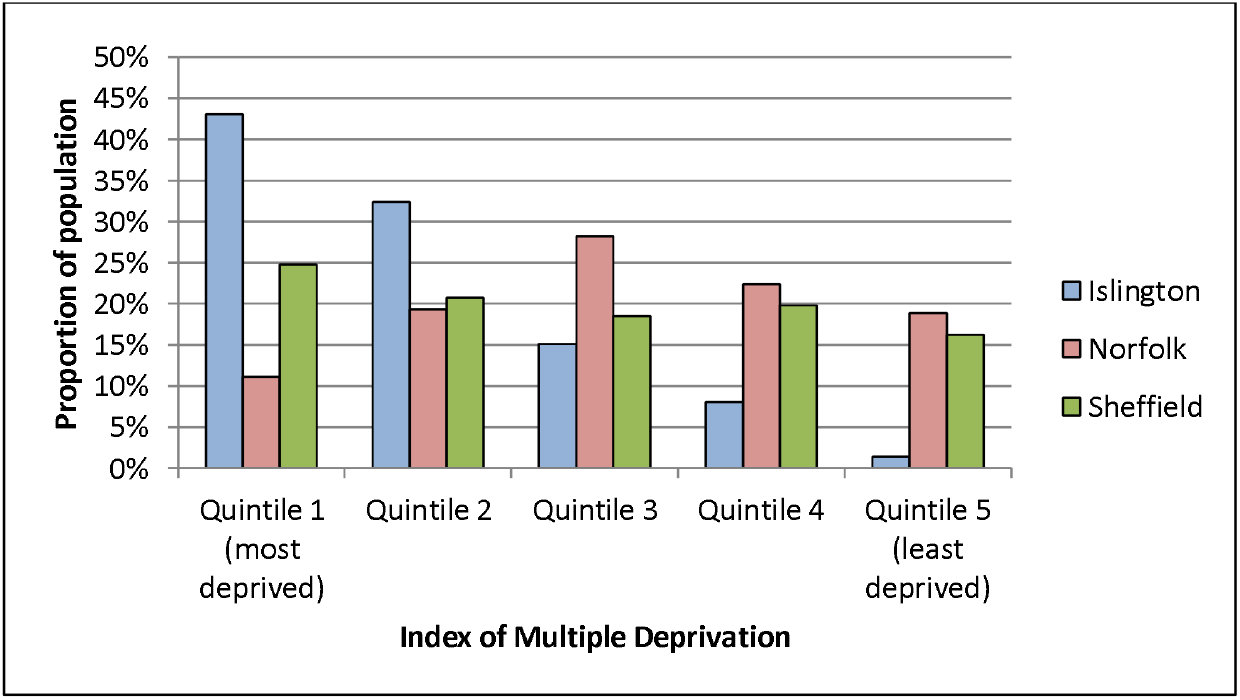
Deprivation distribution of included Local Authorities^1^.

### Measures of availability – Density

Alcohol outlet density measures, also sometimes referred to as ‘container measures’[13] are generally of the form:

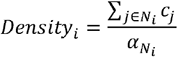

where *i* represents the geographical area of interest, and we are summing the total count of outlets, *c*_*j*_, within a defined neighbourhood, *N*_*i*_, around 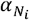 is a weighting factor which represents some characteristic of *N*_*i*_, e.g. population or roadway miles. For this study we defined *N*_*i*_ as a circle of radius 1km around the postcode centroid of *i*, representing a 15 minute walk, and the mean walking journey distance according to the recent National Travel Survey [14]. Our measure is therefore ‘outlet density within 1km’ and as such 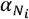 is set to be 1.

### Measures of availability – Proximity

Outlet proximity measures can take several forms, of which the simplest is the minimum distance to the nearest outlet:

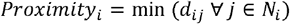

where *d*_*ij*_ represents the distance between *i* and *j*. This distance may be calculated in a variety of ways – here we use the simple Euclidean distance between the postcode centroids of *i* and *j*. As no postcode in mainland England is likely to be a substantial distance from the nearest outlet, we have defined *N*_*i*_ as the Local Authority in question and all bordering Local Authorities.

### Measures of availability – Cluster proximity

Of the measures described above, density measures to not account for the effect of outlets beyond the extent of the defined neighbourhood, *N*_*i*_, while proximity measures consider only the nearest outlet. Cluster measures seek to address these limitations by identifying clusters of outlets which may have a particular draw as a location for a ‘big night out’ for which individuals may travel greater distances than usual. We defined a cluster as any group of four or more on-trade pubs, bars or nightclubs where the maximum distance between any two outlets in the cluster is no more than 250m. Cluster proximity for any point *i* is then given by:

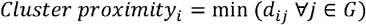

where *G* represents the set of all clusters. Proximity is thus calculated as the distance to the nearest outlet within a cluster.

### Measures of availability – Gravity

Measures of availability or accessibility using gravity or ‘inverse density weighting’ have been used in numerous applications in spatial geography. These seek to calculate the cumulative ‘draw’ on an individual or location of features (alcohol outlets in this case) in the surrounding area and are generally of the form:

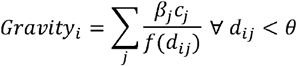

where *β*_*j*_ may be a weighting factor for the outlets at point *j* (e.g. the size or sales volume), *θ* is a maximum distance parameter and *f* is some decay function representing the rate at which the influence of outlets decreases with distance. *θ* is set at 5km based on willingness to travel data for UK grocery stores [15]. A number of specifications for the decay function *f* have been used in the broader spatial geographic literature; the simplest being to assume a linear decay, i.e. that *f*(*d*_*ij*_) = *d*_*ij*_. One limitation of this approach is the issue of ‘self-potential’ – the influence that an outlet has on itself [16,17]. This is an issue with our data as some postcodes contain both outlets and households, which are then allocated the same spatial location (so *d*_*ij*_ = 0 and *Gravity*_*i*_ is undefined). We follow the approach recommended by Owen & Coombes [18] by taking the area of every postcode *Ai*, modelling as a circle of radius 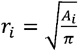 and assuming population density within *i* is uniformly distributed. The mean distance between points (i.e. addresses) within is therefore *0*.*33r*_*i*_, which we use as the value for *d*_*ij*_ where *i* = *j*.

An alternative decay function, as utilised by Branas et al. [9] in their study of alcohol outlets and gun violence, is to set 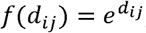, i.e. to assume an exponential decay. This also has the advantage of eliminating the issue of self-potential. As there is no clear theoretical basis to prefer either specification over the other we use both to explore the implications of this choice.

For each of the three Local Authorities, we calculate density, proximity and both gravity measures (linear and exponential decay) for both on- and off-trade outlets separately. Whilst we are not considering the relationship between outlet types and harm outcomes and therefore the type of outlet is not, in itself, important, we do this for illustrative purposes because:

- The patterns of spatial distribution of on- and off-trade outlets are markedly different within England
- On-trade availability is significantly higher than off-trade availability, with over twice as many on- as off-trade outlets in 2014 [19]
- The temporal trends in on- and off-trade outlets are very different, with on-trade outlets decreasing in number by 12% between 2003-14, while off-trade outlet numbers increased by 36% over the same period.

This disaggregation by outlet type therefore gives us a clearer picture of the sensitivity of the different availability measures to differences in spatial distribution, density and temporal trends. For each postcode in Islington, Norfolk and Sheffield we therefore have 9 measures of the availability of alcohol: on- and off-trade outlet density, on- and off-trade outlet proximity, cluster proximity, on- and off-trade gravity using a linear decay model and on- and off-trade gravity using an exponential decay model. All measures are calculated based on outlet location data which includes all neighbouring Local Authorities.

### Statistical analysis

In order to facilitate comparison between measures, they were standardised by converting them to z-scores, i.e. each variable was centered on its mean and divided by its standard deviation to give a measure with mean 0 and standard deviation of 1. As higher values of proximity and cluster distance imply lower availability, while higher values of the density and gravity measures imply lower availability, the proximity measures were inverted around 0 so that higher values equated to greater availability across all measures. Variables were standardised to their 2014 mean and standard deviation to allow standardised temporal trends to be examined. All other analyses use 2014 data only. In addition to descriptive statistics, correlation was assessed using Pearson’s ρ. The extent to which the different measures describe different latent constructs of availability was evaluated using factor analysis.

All statistical analysis was conducted using Stata 12 [20]. Data processing was undertaken in R, with spatial areas calculated using the *rgeos* package [21] and clusters defined using the *dbcan* function in *fpc* [22].Spatial data visualisations were created using QGIS Essen [23].

## Results

### Descriptive statistics

Table 2 presents the mean and interquartile range of the crude (i.e. unstandardized) availability measures across all 3 Local Authorities. This illustrates the high level of availability of alcohol, with an average of 45 outlets within one kilometre of any postcode, and 75% of postcodes having at least 2 on-trade and 1 off-trade outlets within walking distance. Availability is markedly higher in the on- than the off-trade, although nowhere is far from an outlet selling alcohol, with a mean distance of 400m to an on-trade outlet and 900m to an off-trade outlets.

**Table 2.** Summary of measures (unstandardised)

|  |  | Mean availability<br>(Inter-quartile range) |
| --- | --- | --- |
| On-trade | Density (outlets/km) | 34.4 (2-25) |
|  | Gravity (linear) | 136.7 (7.2-111.1) |
|  | Gravity (exponential) | 37.9 (1.8-27.4) |
|  | Proximity (km) | 0.4 (0.1-0.5) |
|  | Cluster proximity (km) | 3.9 (0.6-6.8) |
| Off-trade | Density (outlets/km) | 10.1 (1-11) |
|  | Gravity (linear) | 40 (2.6-45.7) |
|  | Gravity (exponential) | 11 (0.8-12.4) |
|  | Proximity (km) | 0.9 (0.2-1) |

### Temporal trends

Moving to the standardised z-scores to facilitate comparisons between measures, Figures 2 and 3 illustrate the time trends in mean availability across the 3 Local Authorities between 2003 and 2014. For both on- and off-trade we see that density and both gravity measures (linear and exponential decay) show very similar patterns. Cluster proximity (an on-trade only measure) follows a similar trajectory, although the agreement is less clear, while overall proximity is more divergent, particularly in the on-trade, where it describes an opposite trend, with proximity increasing over the period while density and gravity was decreasing. Separating these patterns out by Local Authority and quintiles of deprivation (see Supplementary Material Figures 8-11) displays different underlying patterns, but the same broad conclusion, with density and gravity measures aligning closely while proximity measures describe different and often countervailing trends.

**Figure 2.**
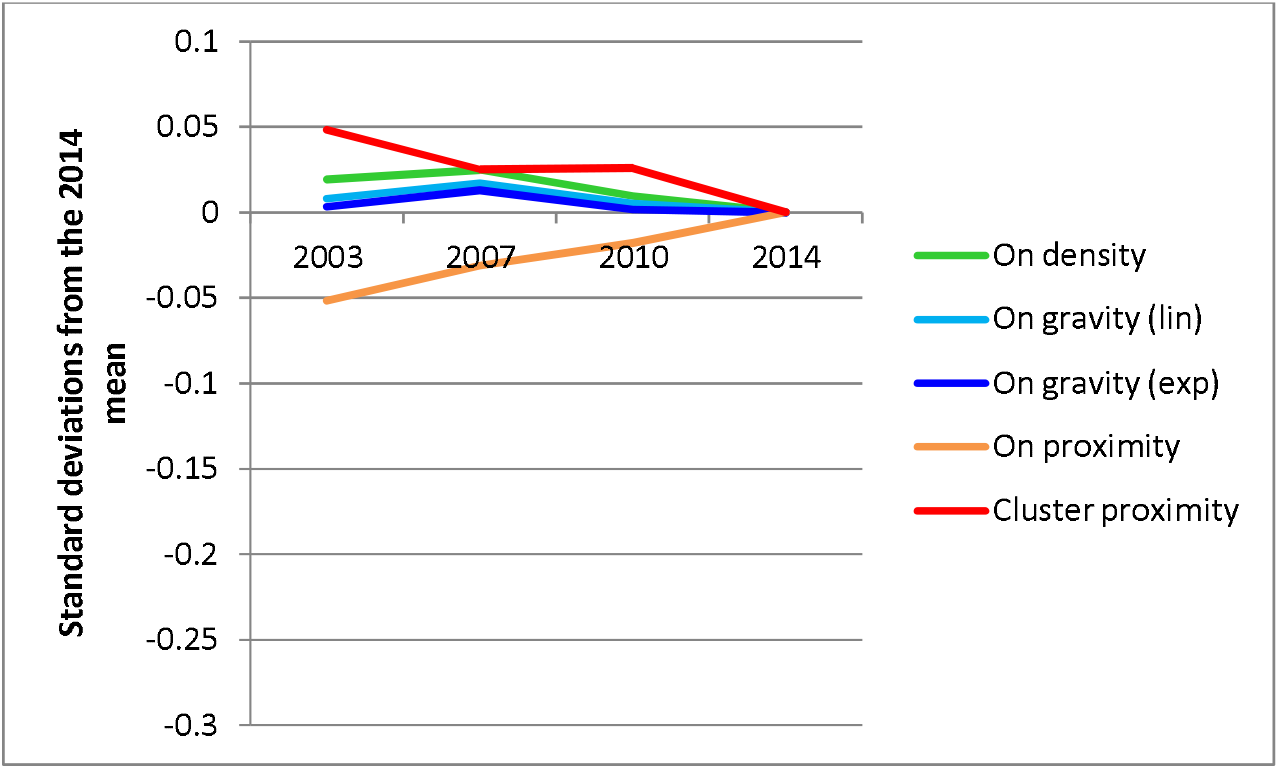
Time trends in on-trade availability.

**Figure 3.**
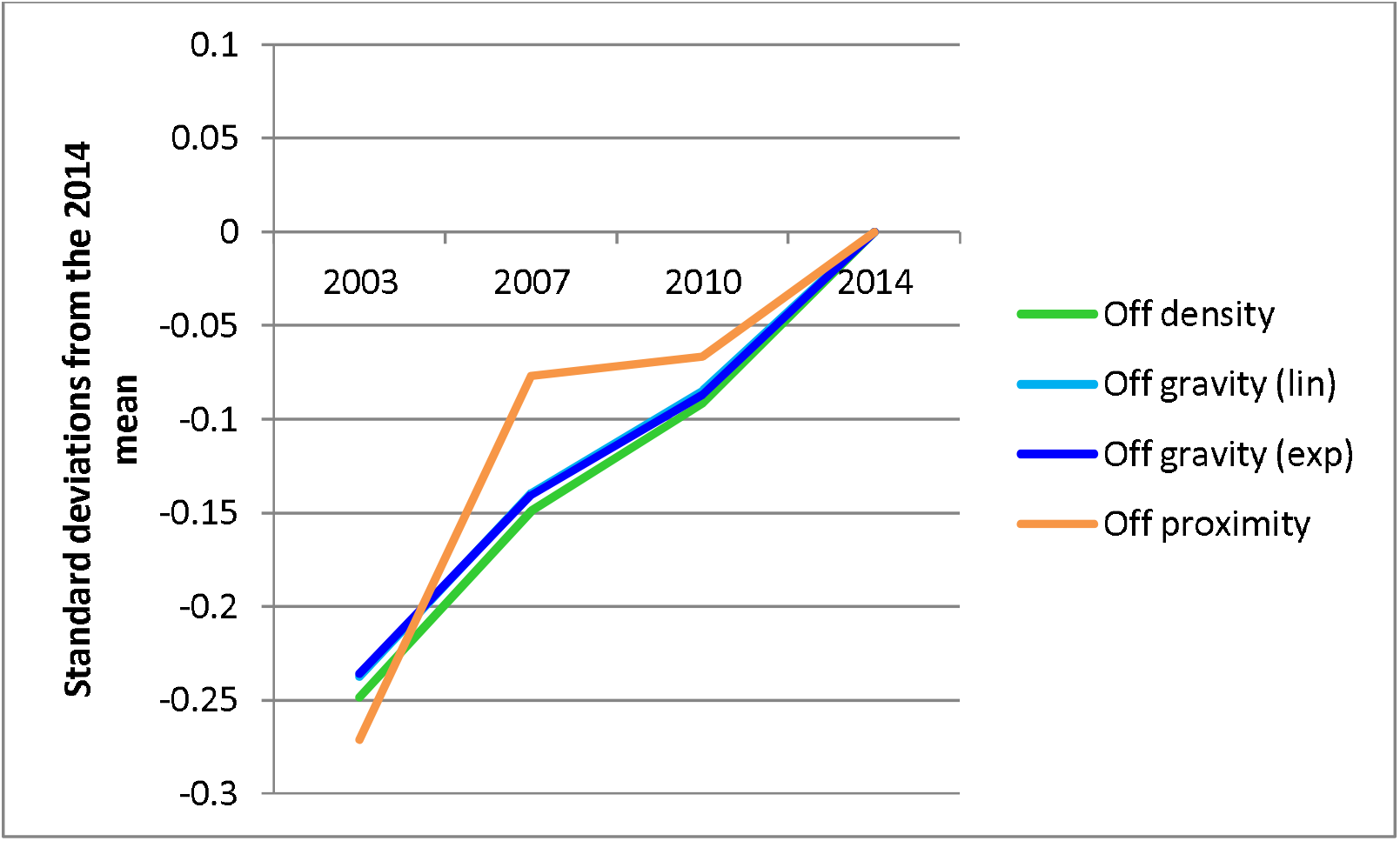
Time trends in off-trade availability.

### Variation by deprivation and Local Authority

In order to explore the extent to which similarities or differences between the calculated measures may be influenced by geographic and socioeconomic factors, Figure 4 illustrates the mean 2014 values for each measure broken down by Local Authority and quintile of deprivation. This shows a number of interesting patterns. Firstly, across all measures, availability is higher in Islington than in Norfolk or Sheffield, although this is much more pronounced for density and gravity than proximity measures. Secondly, across all areas there is close agreement between the density and gravity measures, while the proximity measures show a fundamentally different pattern. Thirdly, the socioeconomic patterning of availability is very different across the three Local Authorities, but broadly similar in both the on- and off-trades within Local Authorities. Under the density and gravity measures, availability is high in the 3 most deprived quintiles in Islington, with the greatest availability in the middle quintile. There is no clear socioeconomic patterning for the proximity measures. In contrast, the greatest availability in Norfolk is in the most deprived quintile for all measures. Availability is highest in the second most deprived quintile in Sheffield according to the density and gravity measures, but again there is no clear pattern for the proximity measures.

**Figure 4.**
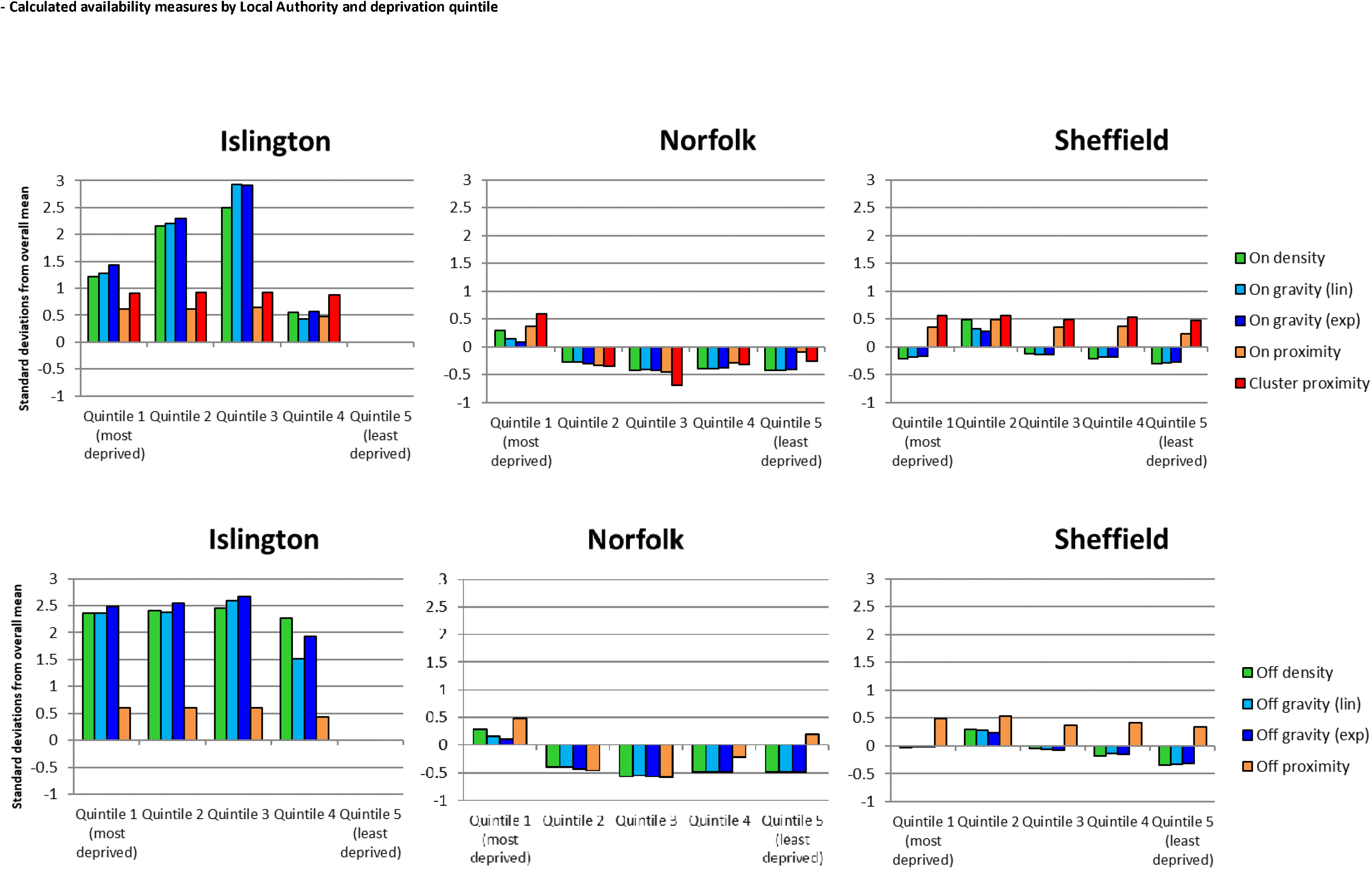
Mean availability by Local Authority and quintiles of deprivation.

### Correlation analysis

Tables 3 and 4 present the correlation matrices for on- and off-trade measures respectively across all 42,036 postcodes in the 3 Local Authorities. As may be expected with such a large sample, all correlations are highly significant (p<0.0001). Both Tables show the same trends as previous results, with strong correlations between the density and both gravity measures, lesser agreement between these and the proximity measures and, in the case of the on-trade, between the two proximity measures themselves. Figures 5 and 6 present these correlations visually, illustrating the strength of the positive relationship between the density and gravity measures and the relatively poor agreement with the proximity measures. Stratifying this analysis by either Local Authority of deprivation quintile (see Supplementary Material Tables 6-9) had no effect on the observed results, although the strength of the intercorrelation between density and gravity measures was somewhat lower in the least deprived areas.

**Table 3.** Correlation matrix for on-trade measures.

| On proximity | On density | On gravity (lin) | On gravity (exp) | Cluster proximity |  |
| --- | --- | --- | --- | --- | --- |
| 1 | 0.2861 | 0.2892 | 0.2777 | 0.4190 | On proximity |
|  | 1 | 0.9635 | 0.9659 | 0.3929 | On density |
|  |  | 1 | 0.9816 | 0.3899 | On gravity (lin) |
|  |  |  | 1 | 0.3915 | On gravity (exp) |
|  |  |  |  | 1 | Cluster proximity |

**Table 4.**
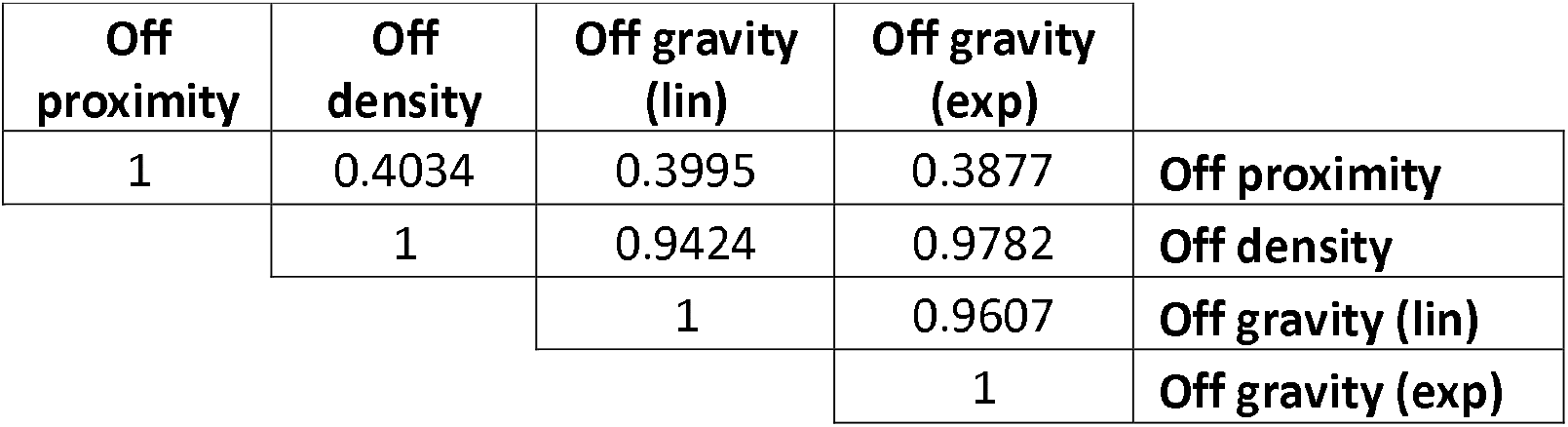
Correlation matrix for off-trade measures.

**Figure 5.**
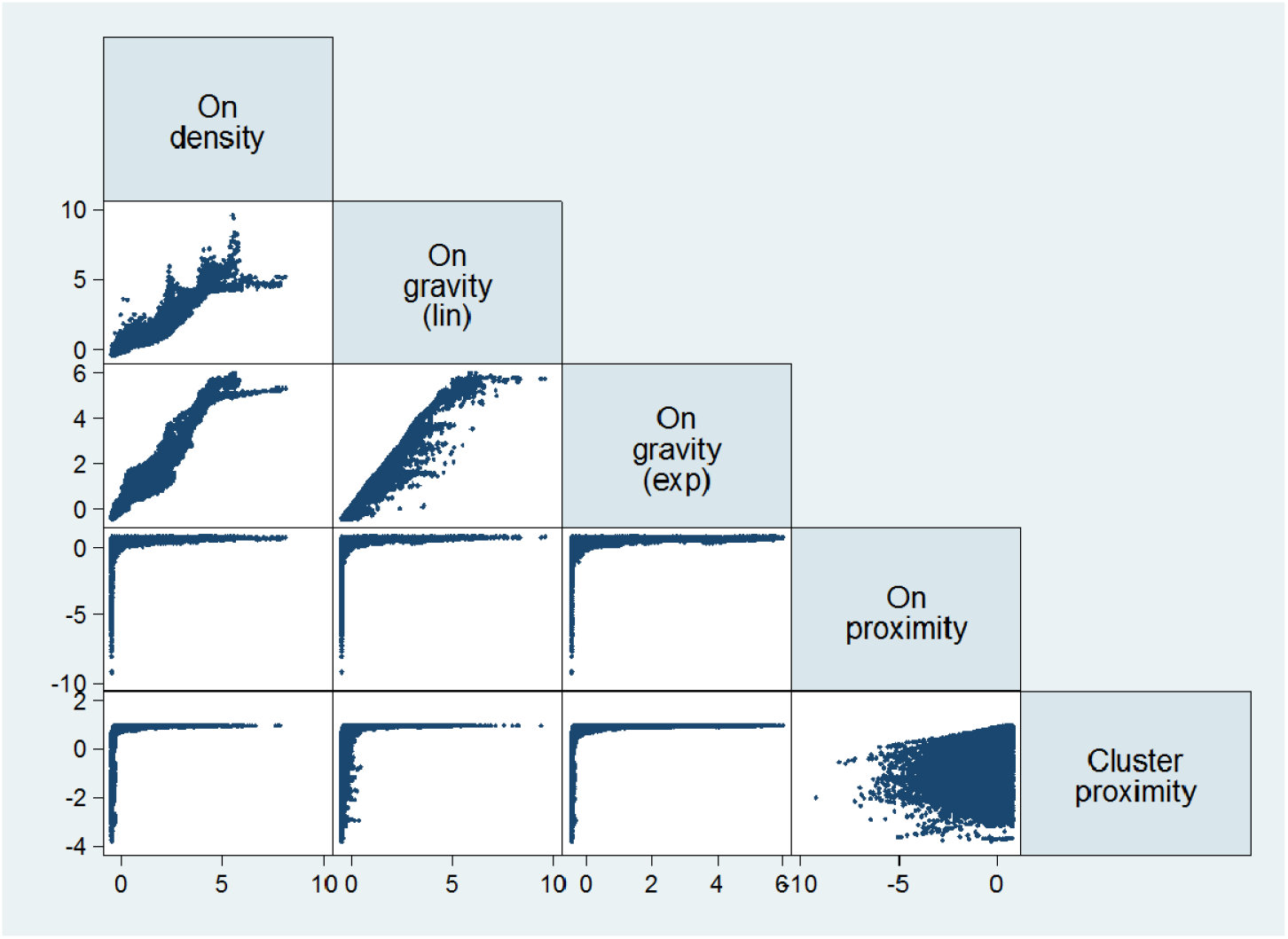
Scatterplot matrix of on-trade availability measures.

**Figure 6.**
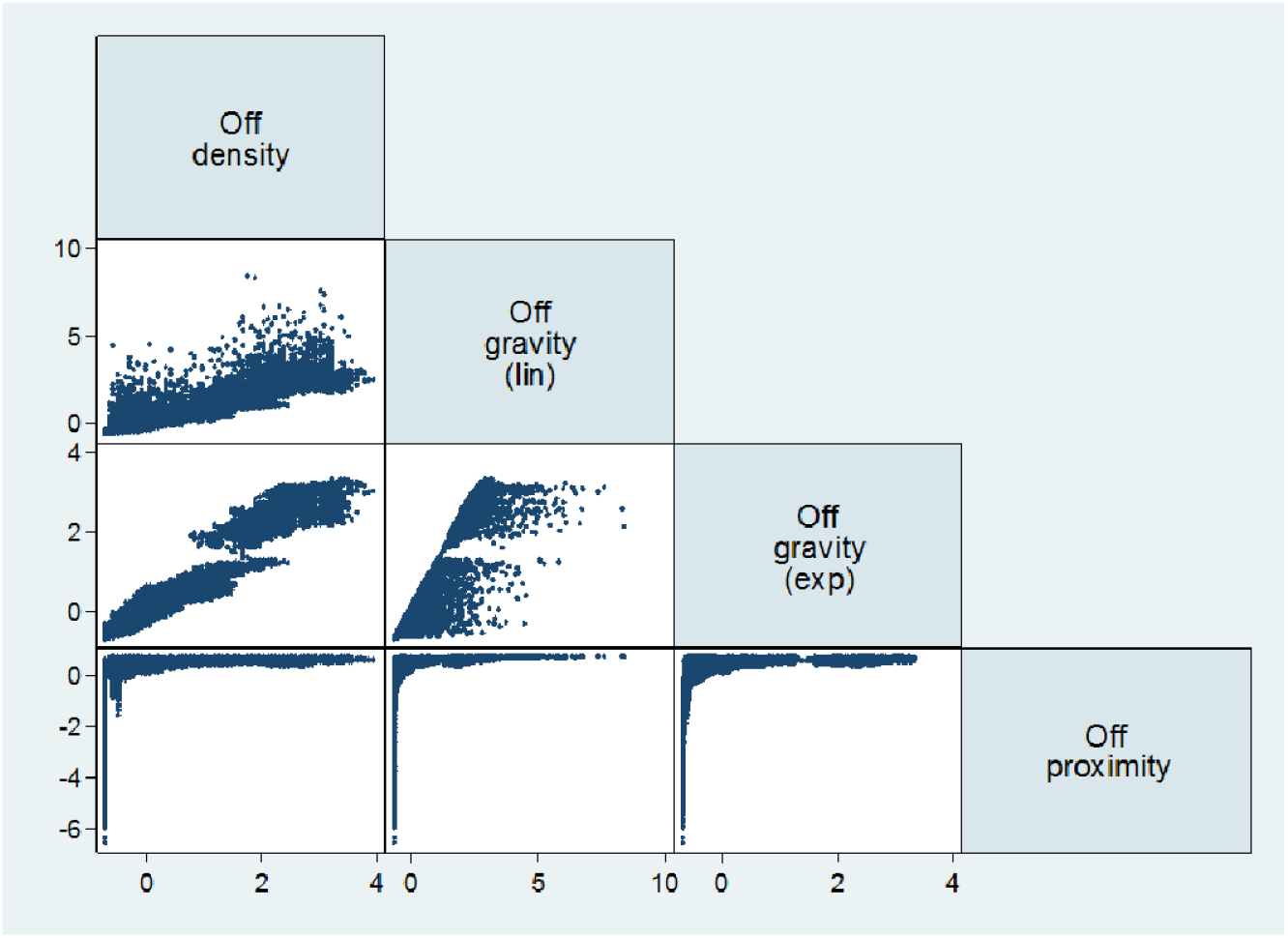
Scatterplot matrix of off-trade availability measures.

### Factor analysis

In order to understand the extent to which the modelled measures may differ because they are capturing different underlying concepts of availability, we applied factor analysis to our measures. The results, presented in Table 5 suggest that the density and gravity measures are primarily describing one factor, which accounts for the vast majority of the variance in results, while the proximity measures are primarily describing a second, lesser factor. This corroborates the findings of both the descriptive and correlation analysis which suggests these two groups of measures are not well aligned.

**Table 5.** Factor analysis results (rotated)

| **On-trade** |  |  |
| --- | --- | --- |
| Factor | Variance | Proportion of variance |
| 1 | 2.99776 | 0.8996 |
| 2 | 0.55479 | 0.1665 |

| Variable | Factor1 | Factor2 | Uniqueness |
| --- | --- | --- | --- |
| On density | 0.9614 | 0.1499 | 0.0533 |
| On gravity (lin) | 0.9773 | 0.1414 | 0.025 |
| On gravity (exp) | 0.9806 | 0.1299 | 0.0215 |
| On proximity | 0.2246 | 0.4936 | 0.7059 |
| Cluster proximity | 0.3262 | 0.5018 | 0.6418 |

| **Off-trade** |  |  |
| --- | --- | --- |
| Factor | Variance | Proportion of variance |
| 1 | 3.01805 | 0.998 |
| 2 | 0.03488 | 0.0115 |

| Variable | Factor1 | Factor2 | Uniqueness |
| --- | --- | --- | --- |
| Off density | 0.9784 | 0.0753 | 0.037 |
| Off gravity (lin) | 0.9599 | 0.0942 | 0.0698 |
| Off gravity (exp) | 0.991 | 0.0168 | 0.0177 |
| Off proximity | 0.3967 | 0.1416 | 0.8226 |

### Spatial visualisations

Finally, in Figure 7, we illustrate 4 of the derived on-trade measures of availability for Islington in order to highlight the geographical differences between the measures which may not be apparent in summary measures already reported (equivalent illustrations for Norfolk and Sheffield can be found in Supplementary Material Figures 12 and 13). This clearly shows the very clear distinction between the density and gravity measures and the proximity measures, but also the subtle distinctions between the former. In particular it is notable that the linear gravity measure uniquely picks out several main roads through the borough, while the density measure identifies a higher-availability area in the north of the borough which is not present in either gravity measure and the exponential gravity measure is dominated by the extremely high number of on-trade outlets to the south of the borough (where the City of London lies).

**Figure 7.**
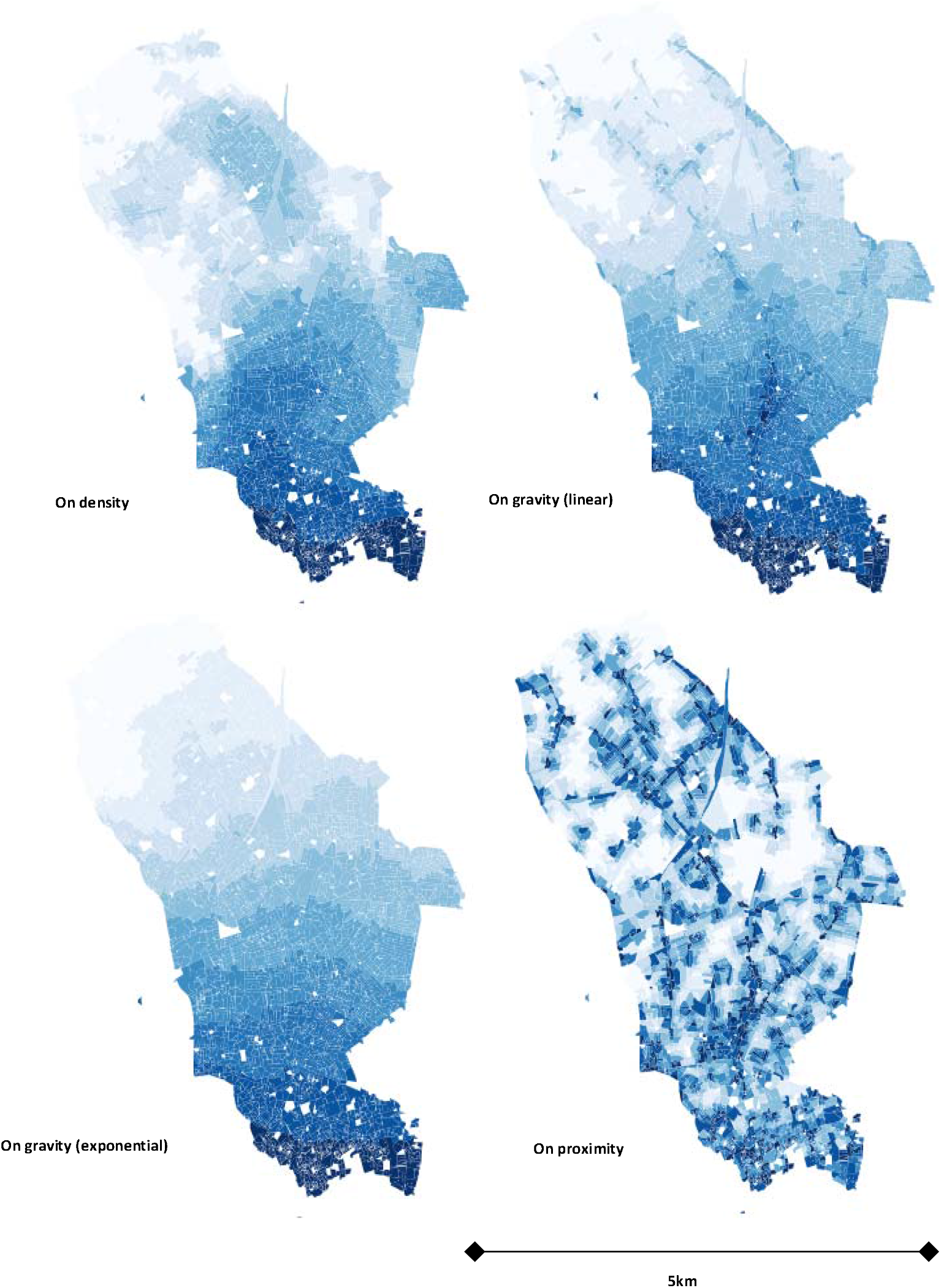
Visualisations of on-trade availability for Islington.

## Discussion

This analysis has demonstrated that there is wide variation in alcohol availability, however it is measured, within the UK, both between, and within, Local Authorities. We have shown that different measures of availability may identify different strengths or even directions of temporal trends. We have identified clear socioeconomic patterning in the distribution of outlets, but this is not consistent across all areas, with the greatest concentration of availability being found in the most deprived, second most deprived and middle quintiles of deprivation in Norfolk, Sheffield and Islington respectively. The strength and nature of this patterning is also affected by the choice of measures, with gravity and density measures showing substantially greater variation than proximity-based measures. We have also shown that the modelled density and gravity measures are very strongly correlated with each other, both longitudinally and cross-sectionally, while proximity measures may not be strongly related either to density and gravity measures or to each other. All results are consistent across both on- and off-trades, suggesting that these conclusions hold across different outlet densities and distributions, at least on the scales found in the UK.

These findings have some significant implications for the study of alcohol availability and its relationship with alcohol-related harm outcomes. Firstly, it is clear that the choice of availability measure can have significant implications for the ways in which availability will be characterised in any study. The use of density or gravity measures may give a very different picture of availability to proximity measures, particularly in areas with large numbers of outlets. The factor analysis presented here suggests that this may be because these different methods are describing different underlying constructs of availability. Secondly, both different availability measures and different geographical areas can give very different pictures of socioeconomic outlet patterning and it may be that observed patterns in outlet distribution may arise from either the choice of measure or be specific to the location of study. These two findings may cast at least some light on recent concerns which have been raised about the consistency of results in availability research [5,6]. They also sound a major cautionary note on the transferability of the findings of availability research between different geographical contexts. If the socioeconomic patterning of availability can vary as widely as suggested in Figure 4 between areas which are no more than a few hundred kilometres apart within the same country, then it is unclear to what extent findings from other countries with different cultures, different alcohol licensing systems and different types of outlet may be relevant. To illustrate this last point with an example: in the UK alcohol is available in almost every local grocery shop or newsagents and therefore the nature of these outlets is not necessarily dominated by their selling alcohol, whereas in the Scandinavian countries the off-trade sale of alcohol is almost exclusively through dedicated state-owned outlets which exist only for that purpose.

There are a number of limitations to this study, primarily that we have only derived and compared measures of availability across 3 Local Authorities in England. Whilst these were purposively chosen to give a spread of sociodemography, it may be that our conclusions would have been different had we chosen a different spread of areas. This study can, however, be regarded as a proof of concept – in finding that both how and where you measure it matter to characterisations of availability across 3 areas we have demonstrated that these are genuine and not just theoretical issues which may affect other studies elsewhere. Another limitation is the restricted set of measures chosen – we did not evaluate alternative ‘container’ measures such as outlets per capita, test alternative density radii than 1km, or different values of the maximum effect range, θ, in the density measures. There are also alternative potential decay specifications (e.g. an inverse square weighting) although we have tested the two most widely used in the broader spatial geographic literature. Finally, as with much of the previous literature, we consider only spatial availability and not temporal or other forms of availability. These limitations should, however, be set against the key strength of the study – the data. We had access to almost uniquely detailed geographic outlet data, allowing us to pinpoint the location of every outlet selling alcohol to within a few metres.

The implications of our findings for future research are clear – different choices of availability measure may lead to very different characterisations of spatial availability and therefore, potentially, very different study results. More attention should therefore be paid to the theoretical understanding of how availability affects behaviour and how different aspects of availability may relate to different harm outcomes. For some outcomes, the minimum travel distance required to purchase alcohol may be the most important factor, for others it may be the cumulative effect of many pubs and bars in a given neighbourhood. There may also be threshold effects in the relationships between measures of availability and outcomes – does the opening of a 20^th^ outlet in an area have the same degree of effect as the 1^st^? Does a reduction in the distance to the nearest off license from 5 to 4km have the same impact as a reduction from 1.1km to 100 metres? There may be a point at which an area becomes ‘saturated’ and further increases in outlet numbers have no meaningful impact on behaviour. Further, different outlet types may have different relationships with behaviour and harm outcomes for different subgroups of the population.

An alternative possibility to address at least some of these issues may be to approach the problem from another angle and, where tenable, specify *a priori* that a given outcome is associated with higher availability of alcohol, then test the strength of relationship between this and a range of alternative measures of availability. The measure which gives the strongest association may then be considered to be the most meaningful characterisation of availability what relates to that particular outcome.

In conclusion: in alcohol availability research, your choice of measure matters. Density and gravity measures and proximity measures appear to be describing fundamentally different concepts of availability and should not be used interchangeably. Thought should be given to the theoretical basis for using any one availability measure over any other in any study and care should be taken when considering the transferability of findings between different geographies and contexts.

## Data Availability

The underlying availability data is commercially sensitive and cannot be shared.

## Supplementary material

**Figure 8.**
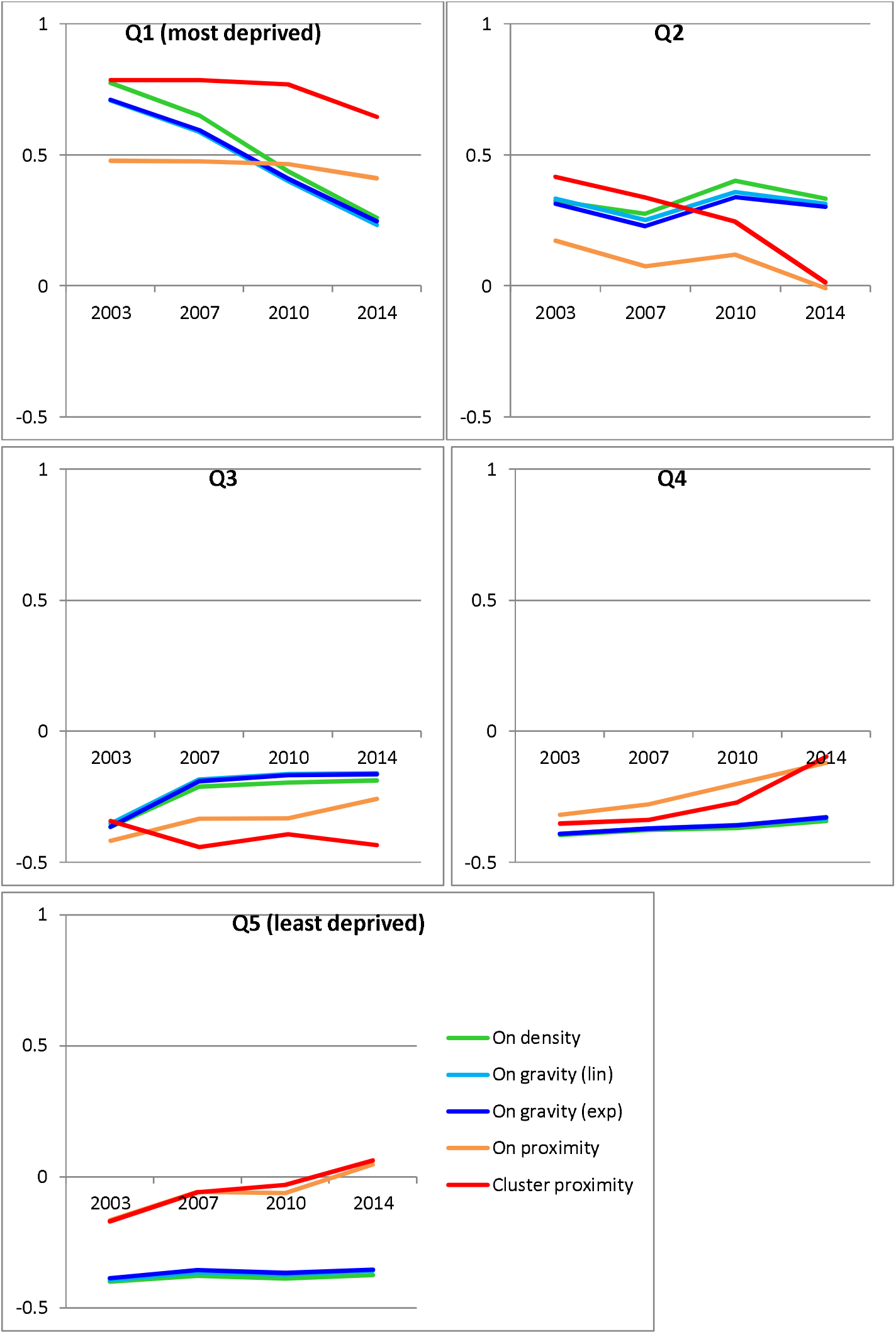
On-trade temporal trends by deprivation quintile.

**Figure 9.**
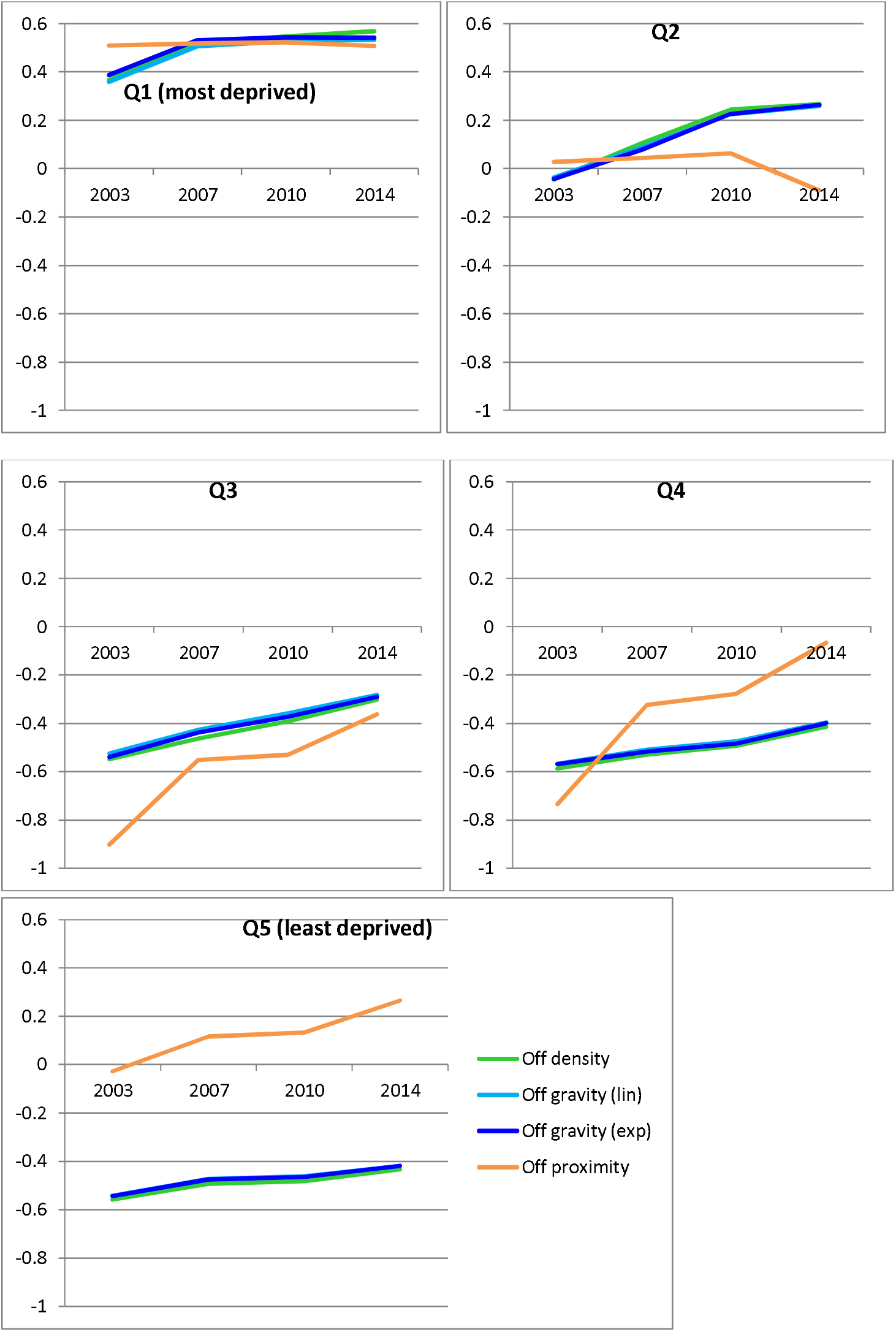
Off-trade temporal trends by deprivation quintile.

**Figure 10.**
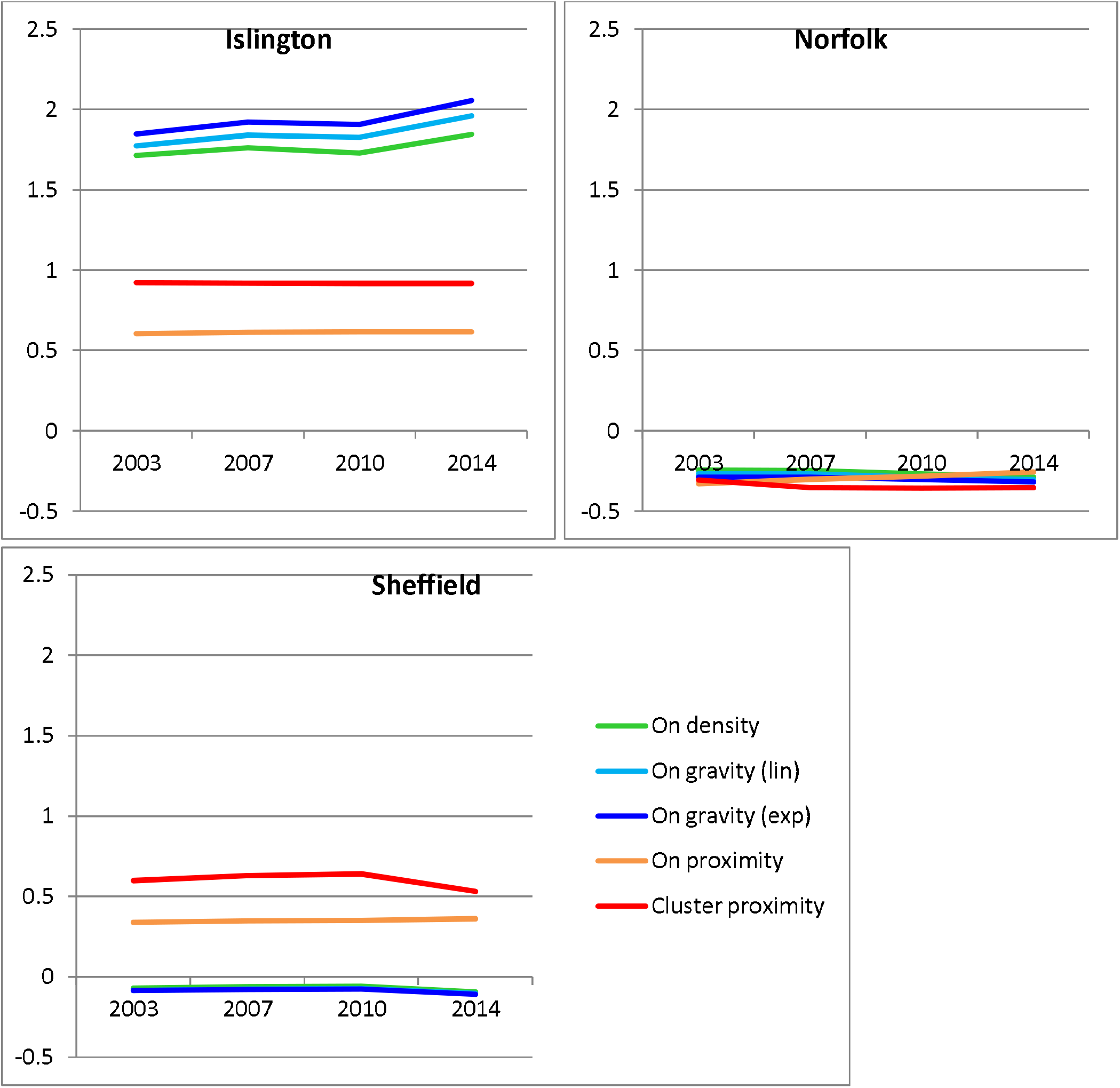
On-trade temporal trends by Local Authority.

**Figure 11.**
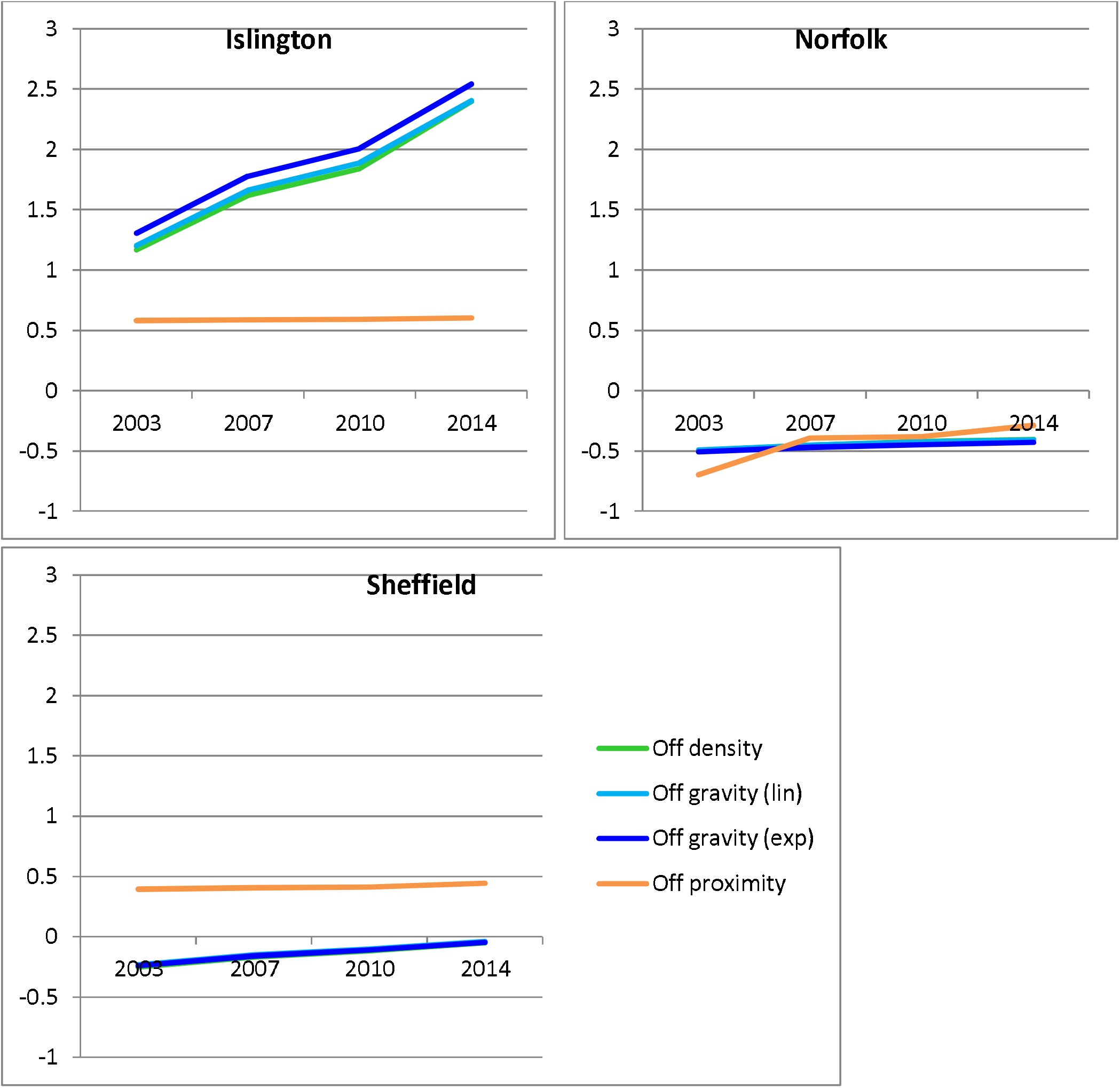
Off-trade temporal trends by Local Authority.

**Table 6.** On-trade measure correlations by deprivation quintile.

**Q1 (most deprived)**
| On proximity | On density | On gravity (lin) | On gravity (exp) | Cluster proximity |  |
| --- | --- | --- | --- | --- | --- |
| 1 | 0.3555 | 0.3695 | 0.3373 | 0.5344 | On proximity |
|  | 1 | 0.9494 | 0.9326 | 0.4401 | On density |
|  |  | 1 | 0.9754 | 0.4498 | On gravity (lin) |
|  |  |  | 1 | 0.4349 | On gravity (exp) |
|  |  |  |  | 1 | Cluster proximity |

**Q2**
| On proximity | On density | On gravity (lin) | On gravity (exp) | Cluster proximity |  |
| --- | --- | --- | --- | --- | --- |
| 1 | 0.3201 | 0.322 | 0.3101 | 0.4324 | On proximity |
|  | 1 | 0.9663 | 0.9727 | 0.4393 | On density |
|  |  | 1 | 0.9787 | 0.4337 | On gravity (lin) |
|  |  |  | 1 | 0.432 | On gravity (exp) |
|  |  |  |  | 1 | Cluster proximity |

**Q3**
| On proximity | On density | On gravity (lin) | On gravity (exp) | Cluster proximity |  |
| --- | --- | --- | --- | --- | --- |
| 1 | 0.2342 | 0.2256 | 0.2163 | 0.3041 | On proximity |
|  | 1 | 0.9766 | 0.9852 | 0.3502 | On density |
|  |  | 1 | 0.9897 | 0.3321 | On gravity (lin) |
|  |  |  | 1 | 0.338 | On gravity (exp) |
|  |  |  |  | 1 | Cluster proximity |

**Q4**
| On proximity | On density | On gravity (lin) | On gravity (exp) | Cluster proximity |  |
| --- | --- | --- | --- | --- | --- |
| 1 | 0.3361 | 0.3565 | 0.3274 | 0.3807 | On proximity |
|  | 1 | 0.9152 | 0.9452 | 0.461 | On density |
|  |  | 1 | 0.9719 | 0.4671 | On gravity (lin) |
|  |  |  | 1 | 0.4763 | On gravity (exp) |
|  |  |  |  | 1 | Cluster proximity |

**Q5 (least deprived)**
| On proximity | On density | On gravity (lin) | On gravity (exp) | Cluster proximity |  |
| --- | --- | --- | --- | --- | --- |
| 1 | 0.3567 | 0.3745 | 0.3396 | 0.2935 | On proximity |
|  | 1 | 0.8827 | 0.9321 | 0.4748 | On density |
|  |  | 1 | 0.9372 | 0.4575 | On gravity (lin) |
|  |  |  | 1 | 0.5095 | On gravity (exp) |
|  |  |  |  | 1 | Cluster proximity |

**Table 7.** Off-trade measure correlation by deprivation quintile.

**Q1 (most deprived)**
| Off proximity | Off density | Off gravity (lin) | Off gravity (exp) |  |
| --- | --- | --- | --- | --- |
| 1 | 0.3199 | 0.3502 | 0.3019 | Off proximity |
|  | 1 | 0.9061 | 0.9598 | Off density |
|  |  | 1 | 0.9404 | Off gravity (lin) |
|  |  |  | 1 | Off gravity (exp) |

**Q2**
| Off proximity | Off density | Off gravity (lin) | Off gravity (exp) |  |
| --- | --- | --- | --- | --- |
| 1 | 0.469 | 0.4662 | 0.4516 | Off proximity |
|  | 1 | 0.9498 | 0.9809 | Off density |
|  |  | 1 | 0.9639 | Off gravity (lin) |
|  |  |  | 1 | Off gravity (exp) |

**Q3**
| Off proximity | Off density | Off gravity (lin) | Off gravity (exp) |  |
| --- | --- | --- | --- | --- |
| 1 | 0.3869 | 0.3751 | 0.3615 | Off proximity |
|  | 1 | 0.951 | 0.9849 | Off density |
|  |  | 1 | 0.9651 | Off gravity (lin) |
|  |  |  | 1 | Off gravity (exp) |

**Q4**
| Off proximity | Off density | Off gravity (lin) | Off gravity (exp) |  |
| --- | --- | --- | --- | --- |
| 1 | 0.4924 | 0.4727 | 0.485 | Off proximity |
|  | 1 | 0.8377 | 0.9514 | Off density |
|  |  | 1 | 0.8885 | Off gravity (lin) |
|  |  |  | 1 | Off gravity (exp) |

**Q5 (least deprived)**
| Off proximity | Off density | Off gravity (lin) | Off gravity (exp) |  |
| --- | --- | --- | --- | --- |
| 1 | 0.4505 | 0.458 | 0.4323 | Off proximity |
|  | 1 | 0.7602 | 0.9146 | Off density |
|  |  | 1 | 0.8263 | Off gravity (lin) |
|  |  |  | 1 | Off gravity (exp) |

**Table 8.** On-trade measure correlations by Local Authority.

**Islington**
| On proximity | On density | On gravity (lin) | On gravity (exp) | Cluster proximity |  |
| --- | --- | --- | --- | --- | --- |
| 1 | 0.3734 | 0.437 | 0.3725 | 0.5055 | On proximity |
|  | 1 | 0.9463 | 0.9662 | 0.4922 | On density |
|  |  | 1 | 0.9747 | 0.5274 | On gravity (lin) |
|  |  |  | 1 | 0.4773 | On gravity (exp) |
|  |  |  |  | 1 | Cluster proximity |

**Norfolk**
| On proximity | On density | On gravity (lin) | On gravity (exp) | Cluster proximity |  |
| --- | --- | --- | --- | --- | --- |
| 1 | 0.3251 | 0.389 | 0.3572 | 0.3438 | On proximity |
|  | 1 | 0.9196 | 0.9434 | 0.3909 | On density |
|  |  | 1 | 0.9533 | 0.4368 | On gravity (lin) |
|  |  |  | 1 | 0.4723 | On gravity (exp) |
|  |  |  |  | 1 | Cluster proximity |

**Sheffield**
| On proximity | On density | On gravity (lin) | On gravity (exp) | Cluster proximity |  |
| --- | --- | --- | --- | --- | --- |
| 1 | 0.2689 | 0.3018 | 0.3014 | 0.3115 | On proximity |
|  | 1 | 0.9405 | 0.9508 | 0.382 | On density |
|  |  | 1 | 0.9506 | 0.4123 | On gravity (lin) |
|  |  |  | 1 | 0.4502 | On gravity (exp) |
|  |  |  |  | 1 | Cluster proximity |

**Table 9.**
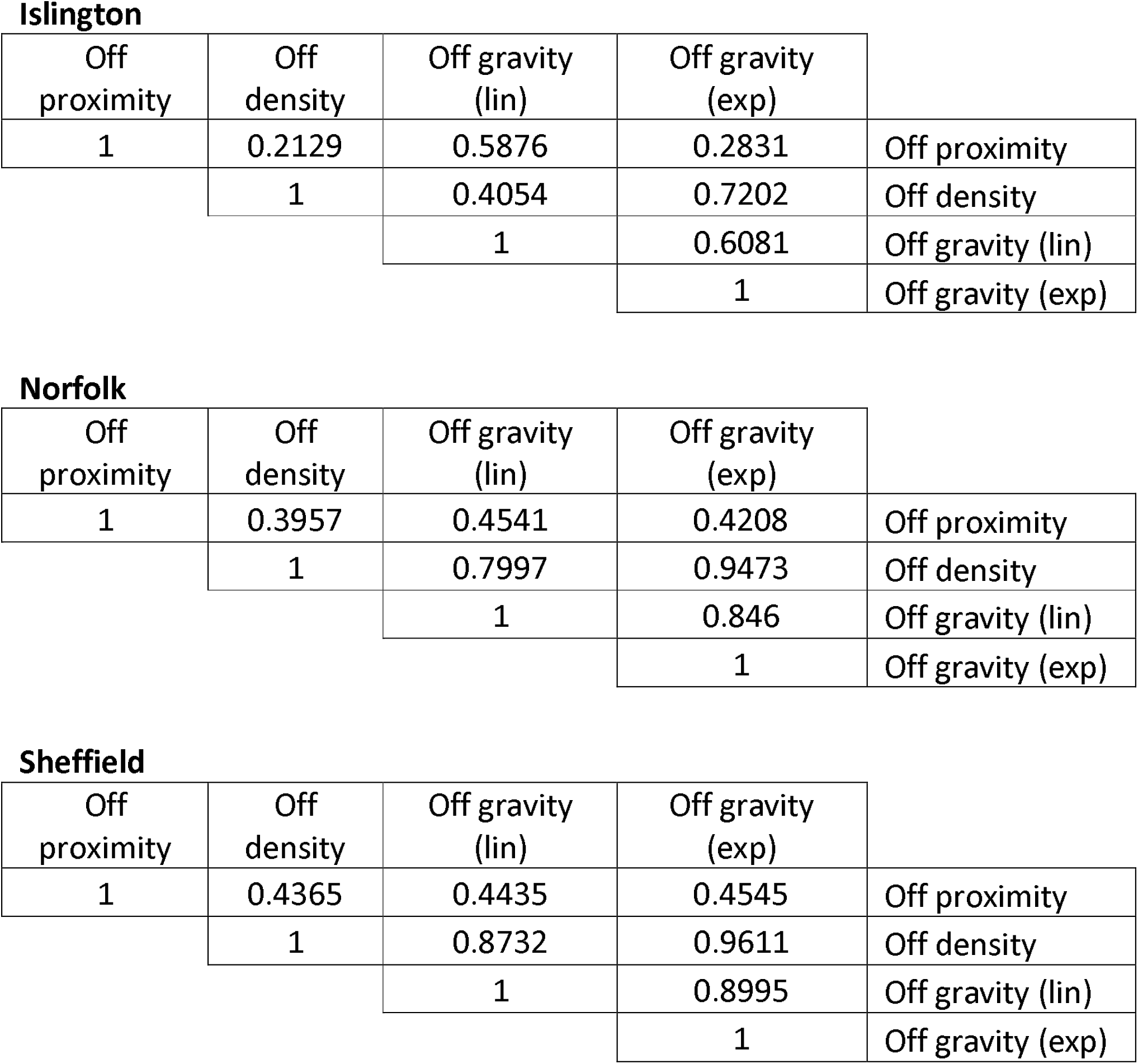
Off-trade measure correlation by Local Authority.

**Figure 12.**
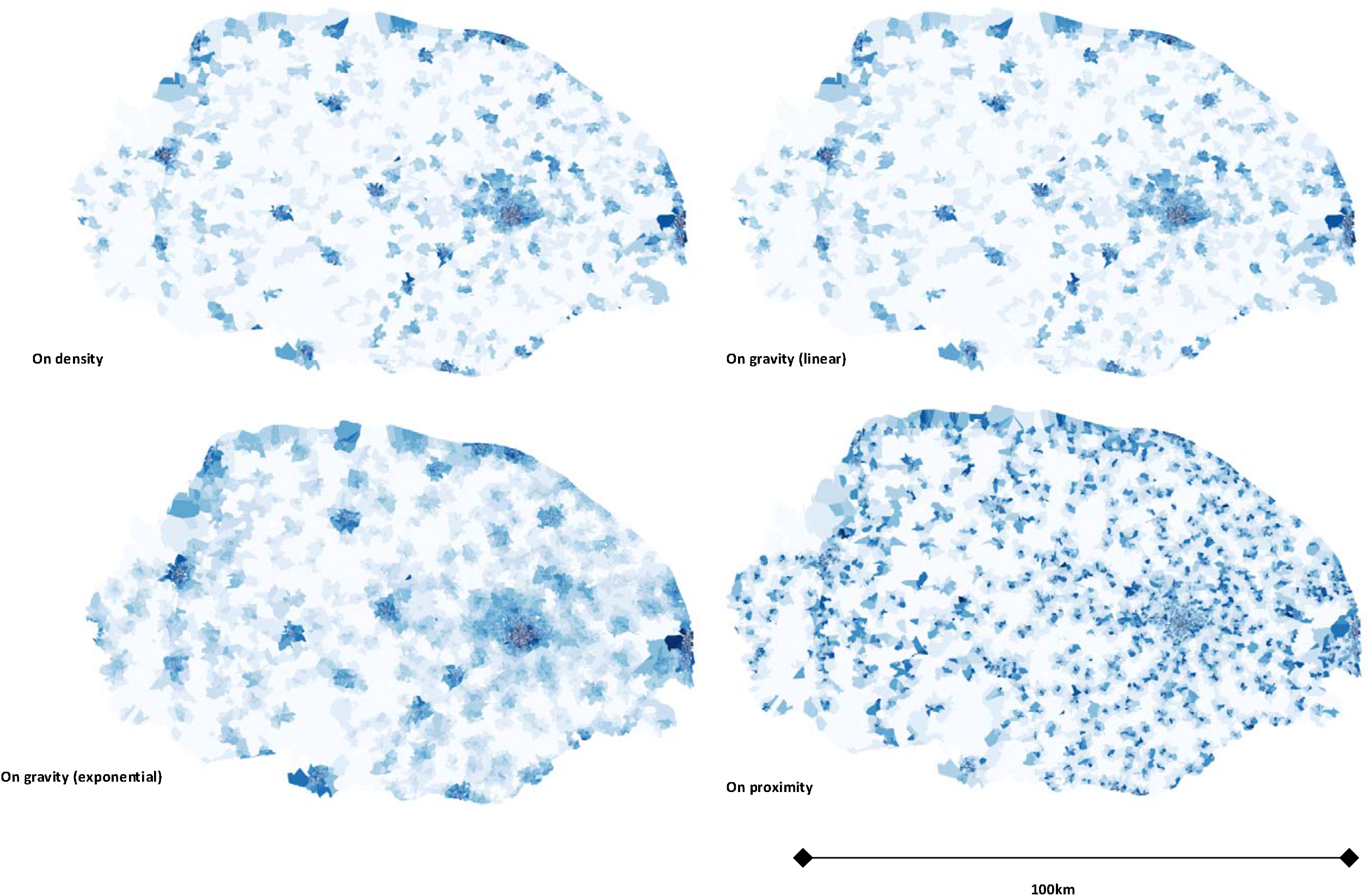
Visualisation of on-trade availability measures for Norfolk.

**Figure 13.**
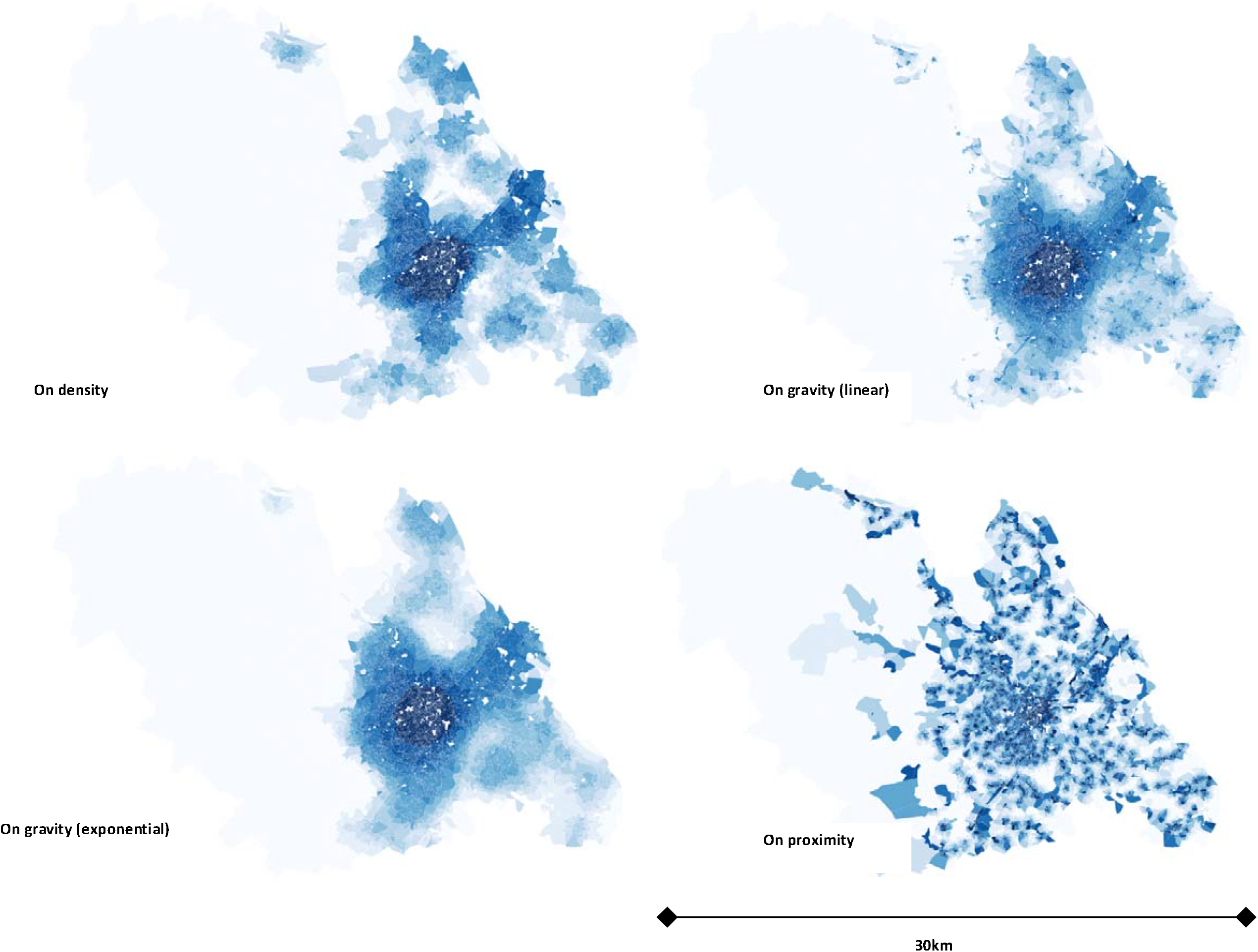
Visualisation of on-trade availability measures for Sheffield.

## Footnotes

1 Quintiles are calculated at the national level (i.e. Quintile 1 represents the 20% most deprived LSOAs in England)

